# Characterization of Oral bacteriome dysbiosis in type 2 diabetic patients

**DOI:** 10.1101/2020.04.09.20052613

**Authors:** Junaid Ahmed Kori, Faizan Saleem, Saeed Ullah, M. Kamran Azim

## Abstract

We aimed to characterize the relationship of the oral microbiome with diabetes in Pakistan. Saliva samples were collected from diabetic patients (n = 49) and healthy individuals (n = 55). 16S metagenomics saliva was carried out by NGS technology. We observed that the phylum Firmicutes (p-value = 0.024 at 95% confidence interval) was significantly more abundant among diabetic patients than among the controls. We found that the abundance of phylum Actinobacteria did not significantly vary among both groups in contrast to a similar report from the USA (Long et al., 2017). On genus level, acidogenic bacteria *Prevotella* (p-value = 0.024) and *Leptotrichia* (p-value = 1.5 × 10^−3^); and aciduric bacteria *Veillonella* (p-value = 0.013) were found to be in higher abundance in diabetic patients. These bacteria are found in dental biofilm and involved in the metabolism of fermentable carbohydrates. Stratified analysis by gender revealed healthy and diabetic females to be more divergent. Abundance of *Prevotella* (p-value = 4.4 × 10-3) and *Leptotrichia* (p-value = 0.015) was significantly associated with male patients. A comparison of oral bacteriome between two groups revealed the dominance of acidogenic and aciduric bacteria in diabetics which suggested the involvement of these eubacteria in oral dysbacteriosis in diabetes mellitus.

## 1. Introduction

Diabetes mellitus is deemed as the most prevalent non-communicable disorder throughout the world with a global prevalence of around 8.3%. It is estimated that ∼387 million people are suffering from this metabolic disorder worldwide (90-95% cases correspond to type 2 diabetes mellitus) [1-3]. It has been asserted that aside from other complications periodontal health is also compromised in type 2 diabetes partly due to alteration in oral microbiome [4].

The human microbiome has been extensively studied in the past few decades because of its dynamicity throughout the body and modulatory potential [5]. The body sites that commonly harbor bacterial communities include oral cavity, skin, vagina, esophagus, colon, etc [6]. Even the secluded and pristine physiological environments (such as nervous system) have recently reported containing bacterial population [7]. A refined estimate based on investigational observations resulted in a ratio of 1.3 bacterial cells per host cell in humans [8].

The term “oral microbiome” has been coined to sum the microbial communities residing in the oral cavity involved in modulation of an array pathophysiological conditions in consortium with each other [8,9]. Bacterial communities in synergetic relationships with each other help to regulate a variety of physiological conditions including pathogen resistance, dysbiosis and immune response [10]. Oral microbiome possesses the capability to transfuse into the bloodstream through the gingival crevice, thus allowing it mobilize throughout the body [11]. Various studies have correlated the oral microbiome with metabolic diseases including dental caries, periodontitis, and gingivitis [12]. An increase in fermentable carbohydrates in the oral cavity or saliva, which is usually the case in diabetes, establishes a favorable environment for the microbes involved in dental caries [13]. The salivary or oral microbiome has been the target of interest for its diagnostic and prognostic value [14]. During the last decade, studies have focused on the management and prevention of certain and disorders such as dental caries by modulating oral microbiome [14, 15].

It has been reported that ∼90% of the oral microbiome cannot be cultured and identified through culturing dependent methods (referred to as “phylotypes” or “uncultivable”). This technical limitation has been a restraint in the identification of a major proportion of oral microbes [16,17]. The application of next-generation DNA sequencing in metagenomics has allowed bypassing these limitations to obtain a considerable microbial profile in a biological system [18]. Periodontal complications have been correlated with variations in the oral microbiome in response to diabetes; however, more data is needed to confer upon such a claim. This study provides additional information on oral bacterial dynamics in type 2 diabetes.

## 2. Materials and Methods

### 2.1 Collection of samples

Before the collection of blood and saliva samples from type 2 diabetic patients and healthy individuals, verbal screening of study participants was carried out. Healthy (non-diabetic) individuals were screened for blood glucose levels and HbA1c according to the authorized guidelines of the American Diabetes Association (ADA). Individuals having un-controlled HbA1c levels (i.e. HbA1c > 6.0), acute or chronic illnesses, periodontal diseases, tobacco usage, pregnancy, and antibiotic usage were not included in the study. Individuals with T2DM according to ADA criteria were selected for the diabetic group. Exclusion criteria for diabetic individuals included current infectious conditions (i.e. white cell count > 11), breastfeeding or pregnancy, medication usage other than anti-diabetics, cancerous malignancies, immunosuppression or immunodeficiency treatment and usage of any form of steroids. According to the exclusion criteria, a total of 104 individuals (i.e. healthy = 55 and diabetics = 49) were selected for the study. Unstimulated saliva samples were obtained from diabetics and healthy individuals from University Clinic, University of Karachi, Karachi, Pakistan and Civil Hospital, Karachi, Pakistan with informed consent. The study was approved by the Independent Ethics Committee (IEC), International Center for Chemical and Biological Sciences, University of Karachi, Karachi, Pakistan.

### 2.2 DNA extraction and quantification

CTAB method was used for metagenomic DNA extraction from saliva samples [20]. Metagenomic DNA quantity was assessed using Qubit 2.0 Fluorimeter (Invitrogen Inc. USA). PCR amplification of the hypervariable V3-V4 region of the 16S rRNA gene was carried according to “16S Metagenomics Sequencing Library Preparation guidelines” (Illumina Inc. USA) by using T100 thermal cycler (BioRad, USA).

### 2.3 Library Preparation, Quantification, Pooling, and 16S rRNA Sequencing

Multiplexing was performed by attaching the dual indices to the PCR amplicons. Briefly, PCR reaction contained PCR amplicon (5 μL), Nextera XT Index Primer 1 (N7XX) (5 μL), Nextera XT Index Primer 2 (S5XX) (5 μL), 2x KAPA HiFi HotStart ReadyMix (25 μL), and volume was adjusted to 50 μL by PCR grade water. The thermal cycling conditions were set as preheating at 95 °C for 3 minutes, 8 cycles of 95 °C for 30 seconds, 55 °C for 30 seconds, 72 °C for 30 seconds followed by a final extension at 72 °C for 5 minutes, and hold at 4 °C. Index PCR clean-up was carried according to the “16S Metagenomics Sequencing Library Preparation” guidelines. Libraries were pooled in equimolar concentration in a single tube. Finally, pooled libraries were sequenced by the MiSeq platform (Illumina Inc., USA) with MiSeq reagent kit V3 (2×300bp).

### 2.4 Bioinformatics and Data analysis

Quality analysis of paired-end NGS reads was carried by FastQC [21]. For quality filtration, Priseq-lite^22^ was used with a minimum length of 75 bases/read, and a minimum, mean quality score of 25. The paired-end reads were assembled by PANDAseq Assembler [23]. The alignment and annotation of 16S rRNA reads were carried against the 16S Microbial Database of NCBI. Annotated NGS reads were then analyzed by MEGAN Metagenome Analyzer [24].

### 2.5 Statistical Analysis

Statistical analysis and heat map plotting was performed by STAMP statistical tool and ClustVis web server, respectively [25,26]. In brief, Welch’s t-test with Storey’s FDR correction method was utilized for the differential abundance analysis of bacteriome in both studied groups with a cut-off p-value of 0.05. PCA plot was constructed according to ANOVA (Analysis of variance) with Storey’s FDR correction at the cut-off p-value of 0.05.

## 3. Results

### 3.1 Clinical data of study participants

We carried out 16S rRNA metagenomic analysis of oral bacteriome of type 2 diabetic (T2D) patients in comparison with healthy individuals residing in Karachi, Pakistan. Unstimulated saliva samples [healthy n=55 (44 males and 11 females); T2D patients n=49 (28 males and 21 females)] were collected with the informed consent of participants. The mean age of healthy individuals and patients was 39.7±11.8 and 53.1±7.9 years respectively. The average BMI was 23.8±3.2 and 25.4±4.3 for healthy and diabetics respectively (Table 1).

**Table 1.**
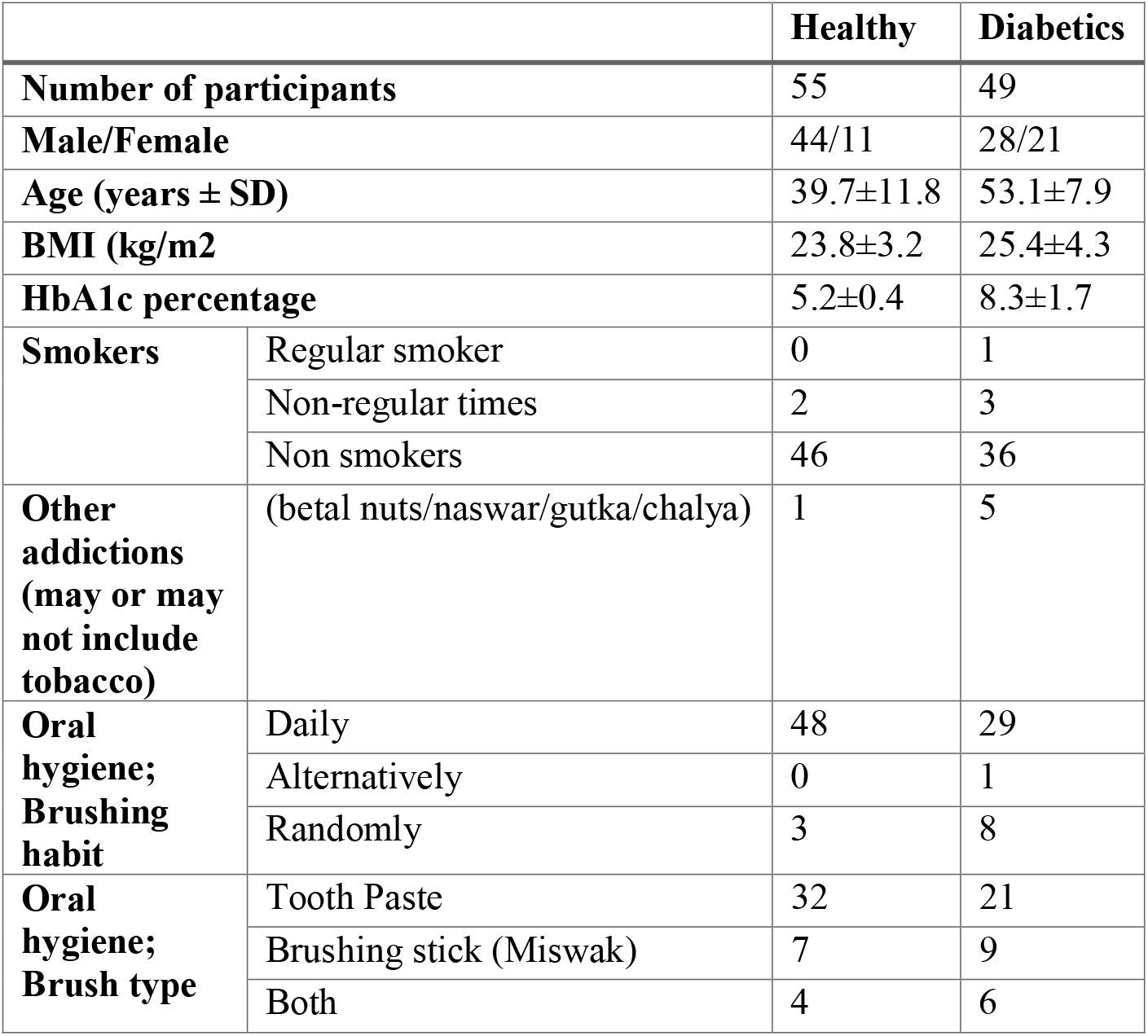
General characteristics and medical profile of healthy (n= 55) and diabetic (n=49) studied individuals.

### 3.2 16S rRNA metagenomic sequencing and alpha diversity analysis

In total, 6,126,515 and 4,254,100 next-generation sequence (NGS) raw reads were obtained for samples from healthy individuals (n=55) and diabetic patients (n=49), respectively. The raw reads were filtered according to quality control parameters, which resulted in 4,175,739 and 2,870,792 reads for healthy individuals and diabetic patients, respectively. The length of filtered sequences was in the range of 250-300 nucleotides. Bioinformatics analysis revealed hundreds of bacterial genera cumulatively. The number of bacterial genera identified and alpha diversity indices for both groups is given in table 2 (alpha diversity indices of each sample are given in supplementary table 1). Alpha diversity represents the variation of bacterial genera within a particular sample and can be calculated using Shannon and Simpson diversity measurement [27]. The average alpha diversity (i.e. Simpson’s) for healthy individuals was 11.0±4.6, while for the diabetic patients it was observed to be 9.99±3.7. Hence, saliva samples from healthy individuals demonstrated slightly higher alpha diversity values in comparison to the samples from diabetic patients.

**Table 2:** Number of genera, alpha diversity in Simpson’s and Shannon-Weaver indices of healthy (n=55) and type 2 diabetics (n=49).

|  |  | Number of bacterial genera identified | Simpson's reciprocal index | Shannon-Weaver index |
| --- | --- | --- | --- | --- |
| **Healthy** | Total | 3,993 | 604.76 | 229.09 |
| | Average | 72.6 | 11.0SD±4.6 | 4.17SD±0.6 |
| **Diabetics** | Total | 3,754 | 489.48 | 199.09 |
| | Average | 77 | 9.99SD±3.7 | 4.06SD±0.5 |

### 3.3 Characterization of salivary bacterial communities

Substantial bacterial diversity was observed amidst the saliva samples from healthy and diabetic individuals. Among the top 10 bacterial phyla (Figure 1), Proteobacteria (Healthy = 36.3%, Diabetics = 28.5%) was predominant in both groups followed by Bacteroidetes (Healthy = 29%, Diabetics = 30.8%), Firmicutes (Healthy = 20.2%, Diabetics = 23.6%), Fusobacteria (Healthy = 6.6%, Diabetics = 7.9%) and Actinobacteria (Healthy = 5.0%, Diabetics = 6.2%). Figure 2 provides the abundance of bacterial phyla in both groups using Welch’s t-test with Storey’s FDR correction with 95% confidence interval. Sequences related to phylum Proteobacteria (p-value = 0.013) were significantly abundant in healthy individuals, while Firmicutes (p-value = 0.024) were significantly higher among diabetic patients. Fusobacteria, Actinobacteria and other less dominant phyla demonstrated statistically non-significant variation among both groups.

**Figure 1.**
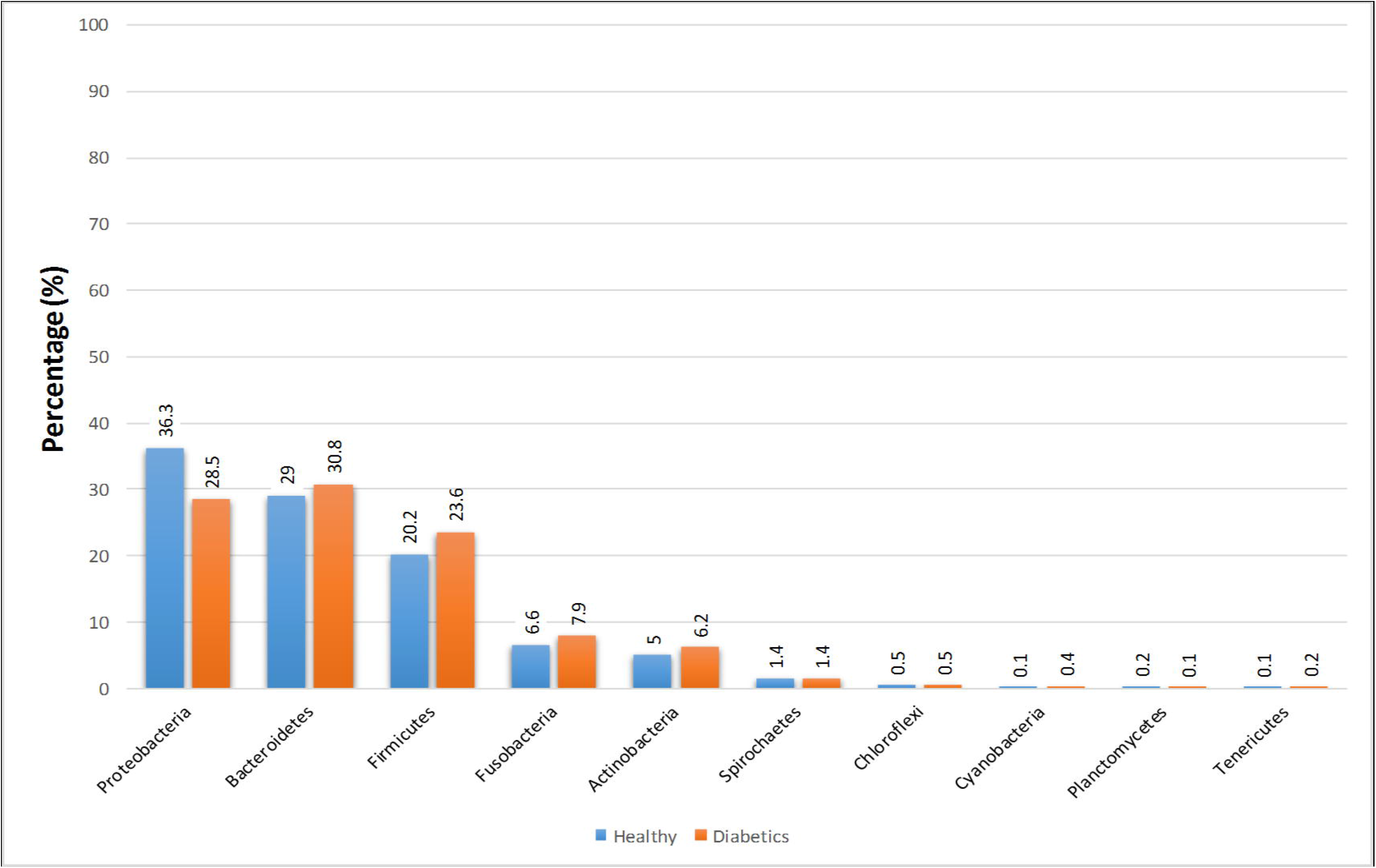
Percent bar plot demonstrating abundance of top 10 bacterial phyla in saliva of healthy (n = 55) and diabetic (n = 49) individuals.

**Figure 2.**
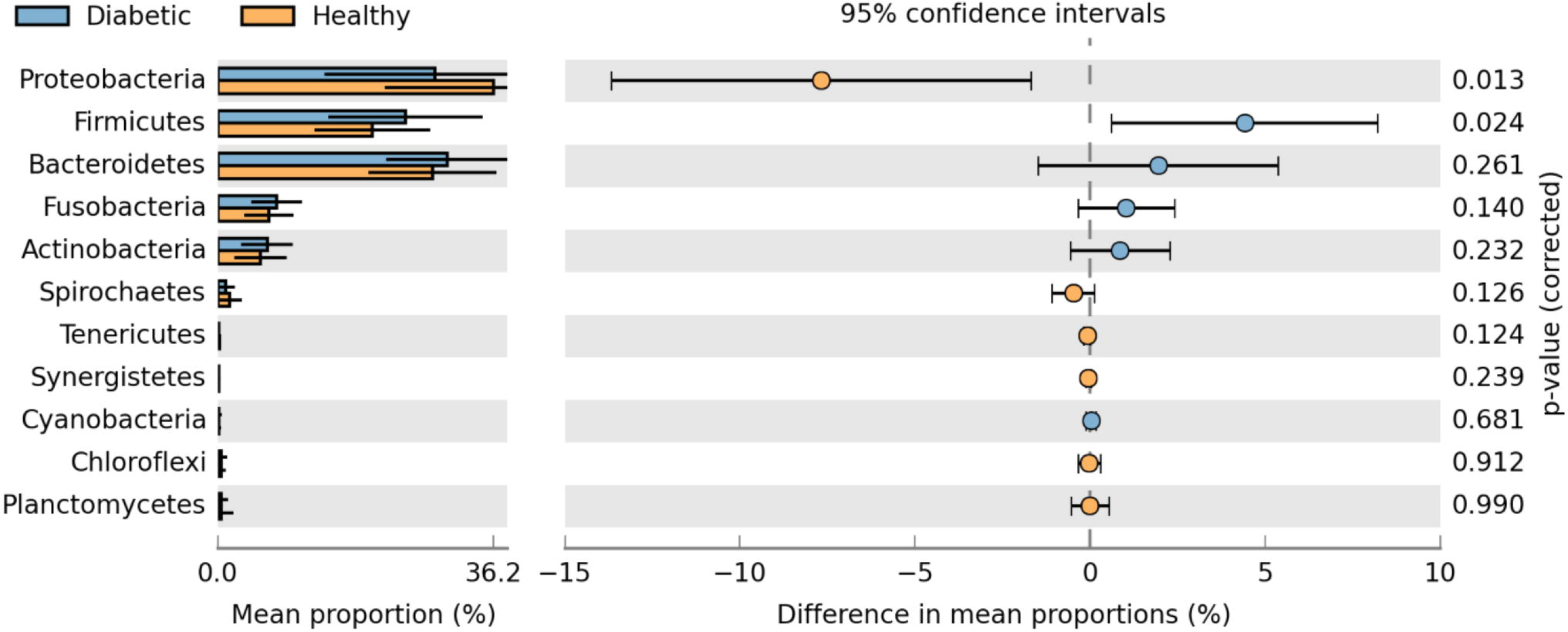
Extended error bar plot representing Welch’s t-test based differential abundance profile of bacterial phyla in healthy (n = 55) and diabetic (n = 49) individuals at 95% confidence interval with Storey’s FDR correction.

The NGS reads from all metagenomic samples were annotated according to bacterial genera. Welch’s exact test with Storey’s FDR correction (p-value of 0.05 and 95% confidence interval) was carried out to assess the salivary bacterial genera whose population significantly increased in response to diabetes (Figure. 3). The analysis of sequence counts showed that the population of eight bacterial genera significantly elevated among diabetic patients. These bacterial genera included *Prevotella* and *Capnocytophaga* (Phylum Bacteroidetes); *Veillonella* (Phylum Firmicutes); *Leptotrichia* (Phylum Fusobacteria). Whereas abundance of *Pseudomonas* (Proteobacteria); Porphyromonas (Bacteroidetes); *Stenotrophomonas* (Proteobacteria) and *Campylobacter* (Proteobacteria) decreased in diabetic patients.

**Figure 3.**
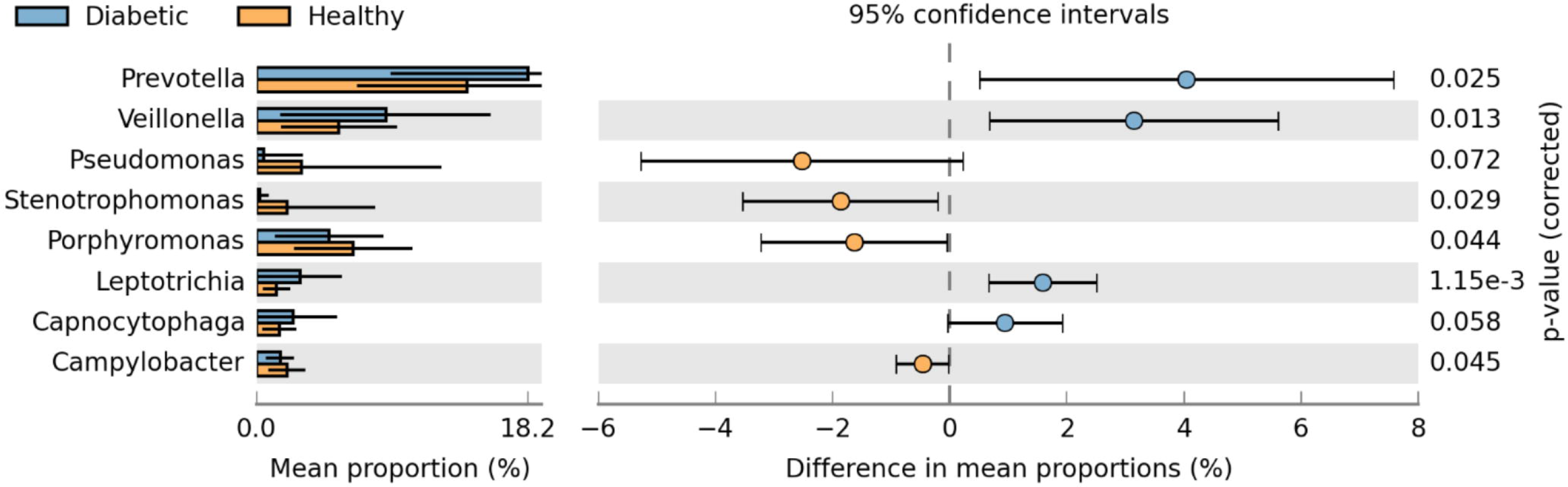
Extended error bar plot representing Welch’s t-test (p-value ≤ 0.05) based differential abundance profile of bacterial genera in healthy (n = 55) and diabetic (n = 49) individuals at 95% confidence interval with Storey’s FDR correction.

Figure 4 illustrates the comparison of the top 40 bacterial genera detected in both studied groups. It was observed that many of the bacterial genera detected in the saliva of healthy individuals were absent in diabetics; while some of the bacterial genera were either exclusive for the saliva of diabetic patients or their population was found to be decreased in healthy individuals. Moreover, the analysis revealed that bacteriome in healthy and diabetic females was more divergent compared to healthy and diabetic males (Figure 4). The similarity threshold PCA plot of healthy and diabetic males resulted in the clustering of the majority of samples in a single area (Figure 5). Figures 6 and 7 demonstrate the gender-based differential abundance profile of healthy and diabetic individuals. The acidogenic bacteriome (i.e. *Prevotella, Veillonella*, and *Leptotrichia*) was higher in male and female diabetics but was less in male and female controls. In male diabetics, sequences related to *Prevotella* (p-value = 0.0044) and *Leptotrichia* (p-value = 0.015) were significantly more abundant. While, in female diabetics, genus *Leptotrichia* was more abundant (with p-value = 0.056).

**Figure 4.**
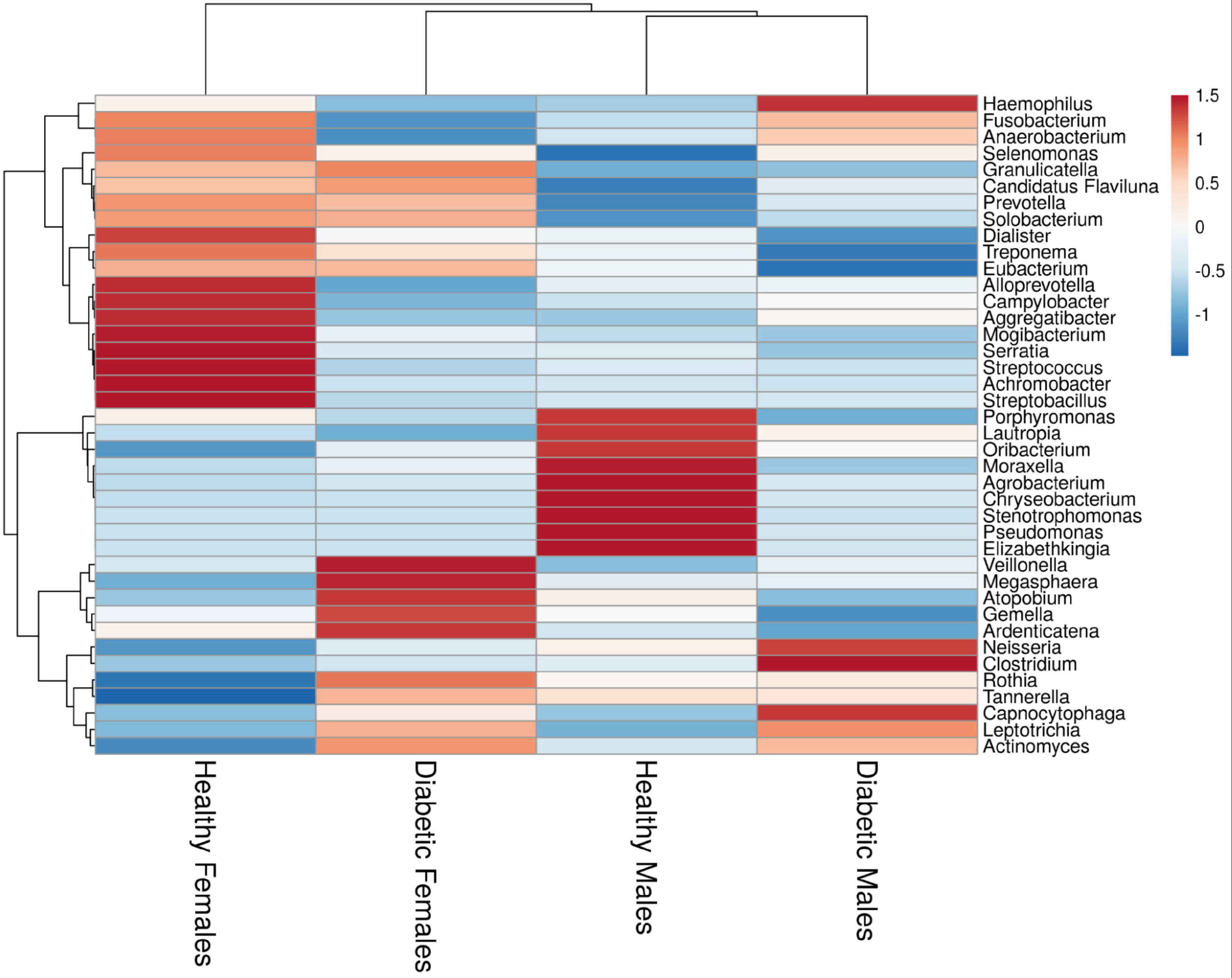
Gender based Heat map plot illustration of top 40 bacterial genera in healthy (n = 55) and diabetic (n = 49) individuals.

**Figure 5.**
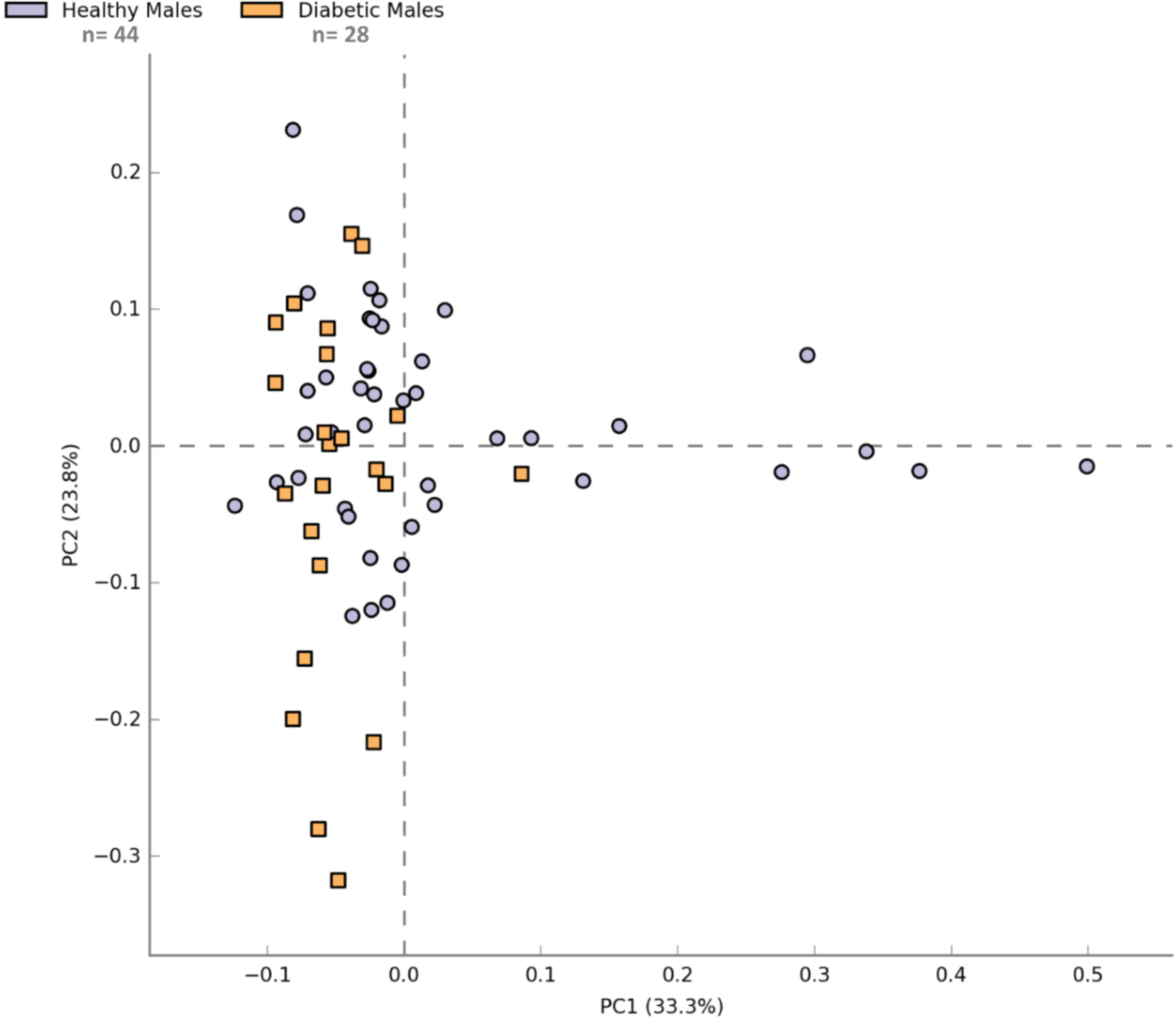
Principle component analysis demonstrating ANOVA (p-value ≤ 0.05) based similarity profile of samples from healthy males (n = 44) and diabetic males (n = 28).

**Figure 6.**
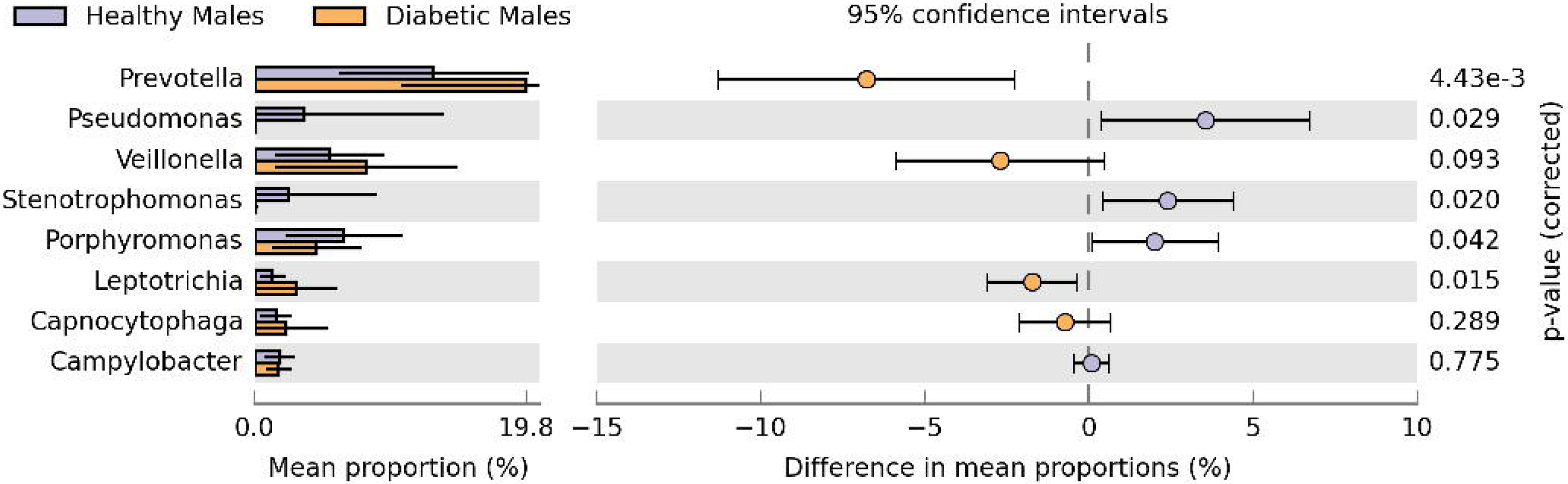
Extended error bar plot representing Welch’s t-test based differential abundance profile of bacterial genera in healthy males (n = 44) and diabetic males (n = 28) individuals at 95% confidence interval with Storey’s FDR correction.

**Figure 7.**
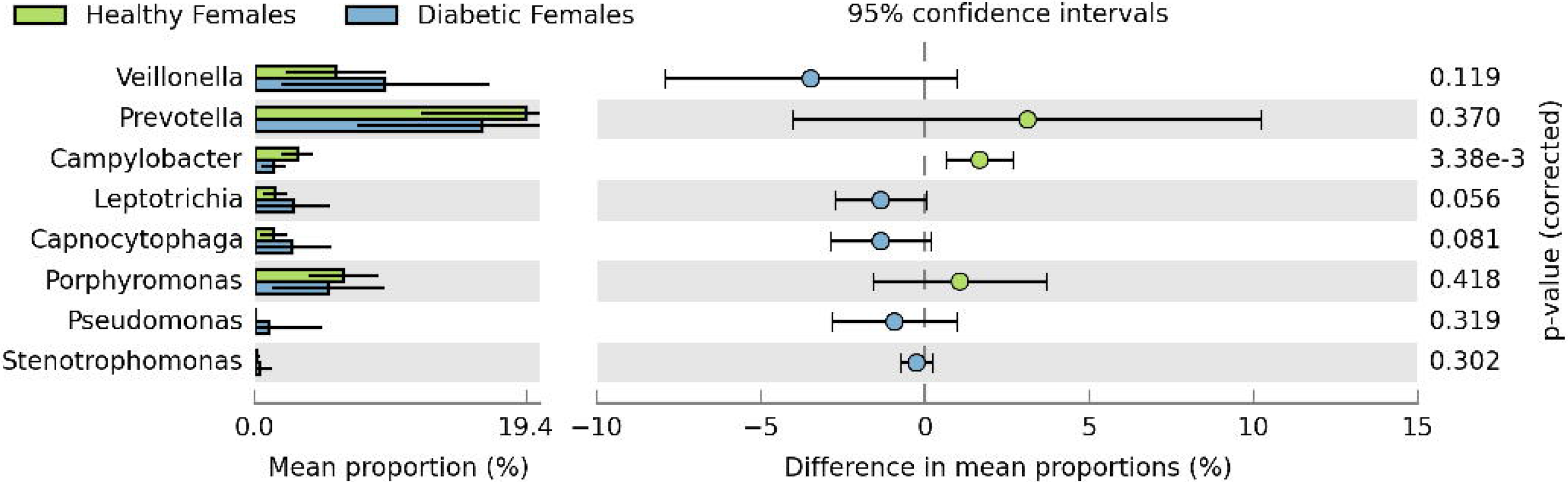
Extended error bar plot representing Welch’s t-test based differential abundance profile of bacterial genera in healthy females (n = 9) and diabetic females (n = 19) individuals at 95% confidence interval with Storey’s FDR correction.

## 4. Discussion

Oral microbiota has been deemed to play a role in the pathogenesis of systemic chronic diseases such as diabetes. It has been estimated that over 700 bacterial species reside in the oral cavity that is involved in the regulation of various immunomodulatory and metabolic processes [28]. Several bacterial species have been reported to be involved in dental caries specifically in response to the progression of diabetes [29]. The present study provides metagenomics based identification of variation in oral bacteriome in patients with diabetes.

In this study, oral microbiota of healthy (n=55) and diabetic (n=49) individuals was analyzed by next-generation sequencing-based metagenomics. The number of bacterial genera identified varied for each studied individual with a higher diversity of oral bacterial genera in healthy individuals. Shannon and Simpson’s diversity indices revealed slightly higher alpha diversity in samples from healthy individuals in comparison to diabetic patients (Supplementary Table 1). Lesser alpha diversity indices in diabetic patients have been attributed to the stressful environmental conditions in the oral cavity due to hyper-mineralization and increased acidouric and/or acidogenic microflora [30].

Evaluation of oral microflora characterized Proteobacteria to be the most abundant bacterial phylum in both groups with a significantly higher population in healthy individuals in comparison to diabetics (Figures 1 and 2). This observation is suggestive of the susceptibility of Proteobacteria in systemic inflammatory metabolic disorders as it has also been described in patients suffering from hepatitis [31].

Moreover, the abundance of Bacteroidetes and Firmicutes was observed to be elevated in the salivary samples of diabetic patients in comparison to healthy individuals. Higher levels of Bacteroidetes have also been characterized in the patients with periodontal diseases [32], while the increase in abundance of Firmicutes has been correlated with higher amount of salivary glucose, which is usually the case for diabetic patients and has been attributed as the favorable environment for the growth of Firmicutes [33]. An increase in inflammatory responses is one of the characteristics of diabetes and the abundance of oral Firmicutes has been correlated with an increase in inflammation [34]. In a study from the USA reported phylum Actinobacteria was present significantly less abundant among the patients with diabetes than among the controls [28]. In contrast, we could not find a statistically significant difference in Actinobacteria among both groups. This population-based difference might be due to variation in lifestyle including dietary habits.

Several bacterial genera were found to be either exclusively or in higher abundance in the saliva of diabetics (Figure 4). Sequences related to genus *Neisseria* were unique to male diabetic patients. It has been proposed in a previous study that this genus is involved in the development of the anaerobic environment in the oral cavity which promotes the growth of nitrate-reducing bacteria which in turn provides a favorable environment for the opportunistic pathogens [35]. Genus *Gemella* that was exclusively present in saliva samples of female diabetics has been reported to be increased in abundance during the onset and progression of inflammatory bowel disease [15]. Genera *Capnocytophaga* and *Actinomyces* were observed to be abundant in both male and female groups which are in agreement with a previous study [36]. Species of genus *Capnocytophaga* are normal inhabitants of the oropharyngeal cavity. Species of this genus have been reported as the causal agents of dental caries and other periodontal diseases in immunocompromised patients [37]. Sequences related to acidogenic bacterial genera including *Prevotella, Veillonella*, and *Leptotrichia* were significantly increased in diabetics compared to healthy individuals (Figure 3). The species of bacterial genera are involved in metabolic utilization of carbohydrates which results in acid production (e.g. lactic acid etc.), thus creating an acidic salivary environment [33]. Species of *Prevotella* have been characterized as the members of oral, vaginal and gut bacterial communities. The abundance of *Prevotella* species occurs in response to an increase in carbohydrate intake [38]. An increase in the abundance of *Prevotella* species has been correlated with periodontal diseases [39]. *Veillonella* species are normal inhabitants of the oral cavity with lactic acid fermentation capability and are correlated with intraradicular infections of apical root canal and dental tubules [40]. *Leptotrichia* species are also members of normal oral flora, which possess lactic acid fermentation ability and characterized as opportunistic pathogens leading to dental caries such as tooth decay [41]. Acidification of the salivary environment in diabetic patients may lead to deterioration of oral and dental health.

The present study provides an overview of variations in oral microflora of diabetic patients in comparison to healthy individuals and might aid in the proficient assessment of complications underlying onset and progression of diabetes.

## Data Availability

The data referred to in the manuscript will be available on request.

